# Contact behaviour before, during and after the COVID-19 pandemic in the Netherlands: evidence from contact surveys in 2016-2017 and 2020-2023

**DOI:** 10.1101/2024.03.06.24303851

**Authors:** Jantien A. Backer, Eric R. A. Vos, Gerco den Hartog, Cheyenne C. E. van Hagen, Hester E. de Melker, Fiona R. M. van der Klis, Jacco Wallinga

## Abstract

**Background:** The first wave of the COVID-19 pandemic in 2020 was largely mitigated by reducing contacts in the general population. In 2022 most contact-reducing measures were lifted.

**Aim:** We assess whether the population has reverted to pre-pandemic contact behaviour and how this would affect the transmission potential of a newly emerging pathogen.

**Methods:** The PIENTER Corona study was held every 2-6 months in the Netherlands from April 2020, as a follow-up on the 2016-2017 PIENTER3 study. In both studies, participants (ages 1-85) reported the number and age group of all face-to-face persons contacted on the previous day. The contact behaviour during and after the COVID-19 pandemic was compared to the pre-pandemic baseline. The transmission potential was examined using the Next Generation Matrix approach.

**Results:** We found an average of 15.4 (14.3-16.4, 95% CI) community contacts per person per day in the post-pandemic period, which is 13% lower than the baseline value of 17.8 (17.0-18.5). Children have the highest number of contacts as before the pandemic. Mainly adults aged 20-59 have not reverted to their pre-pandemic behaviour, possibly because this age group works more often from home. Although the number of contacts is structurally lower compared to the pre-pandemic period, the effect on the transmission potential of a newly emerging respiratory pathogen is limited if all age groups were equally susceptible.

**Conclusion:** Continuous monitoring of contacts can signal changes in contact patterns and can provide a ‘new normal’ baseline. Both aspects are needed to be prepared for a future pandemic.

## Introduction

After the severe acute respiratory syndrome coronavirus 2 (SARS-CoV-2) emerged at the end of 2019, the World Health Organization declared the outbreak a global pandemic on 11 March 2020 [1]. Most nations implemented stringent non-pharmaceutical interventions to mitigate the virus spread, encompassing physical distancing aimed at reducing exposure to SARS-CoV-2 by lowering contact rates. Face-to-face and physical contacts between members of different households were to be reduced, as each contact could be an at-risk event for transmission. Many contact surveys that were conducted during the pandemic showed that more stringent COVID-19 measures indeed lead to lower contact rates [2–10].

During the COVID-19 pandemic, the change in behaviour varied among different groups in the population, either because they were differently affected by control measures or because they differed in ability or willingness to comply. For instance, contacts in younger age groups were greatly affected by school closures [11], while contacts in older age groups decreased due to suspended social gatherings [8,12]. Persons with comorbidities may have reduced their contacts more than persons without comorbidities [8,13,14], given their higher probability of severe outcomes [15]. Also the socio-economic status of a person may have influenced contact behaviour, as persons with a low socio-economic status often have professions that preclude working from home [16].

Most contact-reducing measures were lifted shortly after the emergence of the more transmissible Omicron variants of SARS-CoV-2 in late 2021. The question arises whether contacts in the population reverted to their pre-pandemic levels and how this would affect a future pandemic. Answering these questions would require a contact study in the post-pandemic period, including all age groups, and a valid baseline measurement before the pandemic started. The PIENTER Corona (PiCo) contact study conducted in the Netherlands [17] fits these requirements. The study’s design closely mirrors that of the PIENTER3 study (referred to as ‘the baseline survey’) conducted in 2016-2017 [18], providing the pre-pandemic baseline. Both studies were part of a larger serosurveillance study encompassing a large nationwide sample of the Dutch population. The PiCo survey was conducted every 2-6 months since April 2020 and will continue until the end of 2024. An earlier analysis revealed a reduction of 76% and 41% in the number of contacts outside households during the April 2020 and June 2020 survey rounds, respectively, in comparison to the baseline survey [19].

On 25 February 2022 all contact-reducing measures were lifted in the Netherlands. We aim to compare the contact behaviour in the PiCo survey rounds before and after this date to the contact behaviour before the pandemic. We study the differences in contacts between age groups, medical risk groups and socio-economic groups, and we assess the potential implications for a future outbreak of a respiratory pathogen.

## Methods

### Study design and data

The baseline survey was conducted from February 2016 to October 2017 in a representative sample of the Dutch population [18], who were randomly selected from the national Personal Records Database (BRP) [20]. 80% of these participants were invited for the first round of the PiCo survey conducted in April 2020 [17], and they were reinvited for each subsequent round. The study population was supplemented with a random selection from persons in the BRP older than one year in PiCo rounds 2 (June 2020) [21] and 6 (November 2021) [22], who were also reinvited for each subsequent round. For comparability the baseline participants younger than one year old were excluded from the analysis.

The questionnaires were filled out either by the participant or with help from their parent or guardian if they were younger than 15 years. Participants reported their age and sex, as well as the age and sex of their household members. From a list of self-reported medical conditions, we determined whether participants would be eligible for influenza vaccination according to the national guidelines [23]. Participants with an indication for influenza vaccination were classified as having a high medical risk status.

Participants provided their highest obtained or current education level, that served as a proxy for socio-economic status. The education level was classified according to Dutch standards [24] as low (no education or primary education), medium (secondary school or vocational training), or high (bachelor’s degree, university). Participants up to 14 years old were assigned the highest education level of their parents or guardians. Participants were also asked whether they worked from home in the previous week.

The questionnaire included a section on contact behaviour, where contacts were defined as unique persons with whom the participant talked face-to-face, touched, kissed or played sports. Participants listed the number of contacts they had with persons outside their household on the previous day. The contacts were stratified by age group and either by gender (male/female) or by proximity (less/more than 1.5 meters apart). Participants who failed to provide valid contact data were excluded from the analysis.

A detailed description on the participant selection and participation, the definition of the medical risk status, the validity of contact data, and handling of changes in survey questions is provided in Supplement S1. The data was reformatted to the standard format for contact surveys on socialcontactdata.org and published online [25].

### Contact analysis

The age groups of participants and contacts were aggregated in 10 age groups: 0–4, 5–9, 10–19, 20–29, 30–39, 40–49, 50–59, 60–69, 70–79, and ≥ 80. In each round, the number of community contacts per participant per contact age group was truncated at 50, i.e., a maximum of 500 community contacts per participant. The household composition of the participant was used as a proxy for contacts within the household, assuming the participant would have contacted each household member on the survey day. The community and household contacts were analysed separately.

Within each participant age group, participants were weighted according to the age and sex distribution of the Dutch population [26]. Participants were also weighted according to weekday (weight 5/7) or weekend (weight 2/7), but only for community contacts, assuming the household composition is constant over the week. With these sample weights the weighted mean number of contacts per participant in that participant age group and contact age group was calculated. These numbers are the elements of a contact matrix. Assuming all contacts are reciprocal, we corrected the contact matrix for reporting errors using the age distribution of the Dutch population [27]. The uncertainty was expressed by the 95% bias-corrected bootstrap interval [28], based on 1000 bootstrap samples by participant age group.

The number of community contacts per participant per day was compared to the baseline values that were obtained from the 2016-2017 contact survey. We studied the number of contacts over time and by age group. The reported contacts by medical risk group and education level were expected to be confounded by the age of the participants. To be able to compare the subgroups, we weighted their age groups by the age distribution of the general population and calculated the population average. Such a population average of for instance the high medical risk group should therefore be interpreted as the population average if the entire population would have a high medical risk.

### Analysis of Next Generation Matrix

A total contact matrix was constructed by summing the contact matrices of the community and household contacts. If all age groups were equally infectious and susceptible to a newly emerging pathogen, this matrix can be interpreted as the Next Generation Matrix (NGM) of an epidemic process [29]. In a fully susceptible population without any control measures, the spectral radius (i.e., maximum eigenvalue) of the NGM is proportional to the basic reproduction number R_0_, indicating the average number of secondary cases infected by a typical primary case [30]. The spectral radius of the NGM can therefore be interpreted as a proxy for the transmission potential of a newly emerging pathogen for which no prior immunity exists. We compared the spectral radius over time to the baseline value. The rounds without any control measures (i.e., after round 7) reflect the transmission potential of a newly emerging pathogen under ‘new normal’ circumstances compared to the pre-COVID-19 period. When age groups differ in susceptibility to infection or infectiousness after infection, the NGM changes accordingly. In the early phase of the pandemic, Zhang et al., [2] found that 0-14 year olds are 2.9 times less susceptible than adults aged 15-64 and that adults of 65 and older are 47% more susceptible than adults aged 15-64. We used these values to determine the transmission potential of a newly emerging pathogen that would resemble the early SARS-CoV-2 virus.

## Results

### Study population

The final data set comprises 62490 questionnaires, filled out by 13826 unique participants, 97% of which also provided their household composition. The number of participants in the PiCo survey ranged from 2594 in the first round when only baseline participants were reinvited to 8144 in the sixth round when the study population was supplemented (Tab. **1**). In total 5768 persons participated in the baseline survey (round 0 in Tab. **1**), 2487 of who also participated in one or more (on average 6.6) rounds of the PiCo survey. The participants recruited in round 2 and 6 participated in on average 6.1 and 3.4 rounds. From the baseline and PiCo surveys 387 participants younger than one year were excluded, and 1780 questionnaires without valid contact data were already excluded before.

**Table 1:**
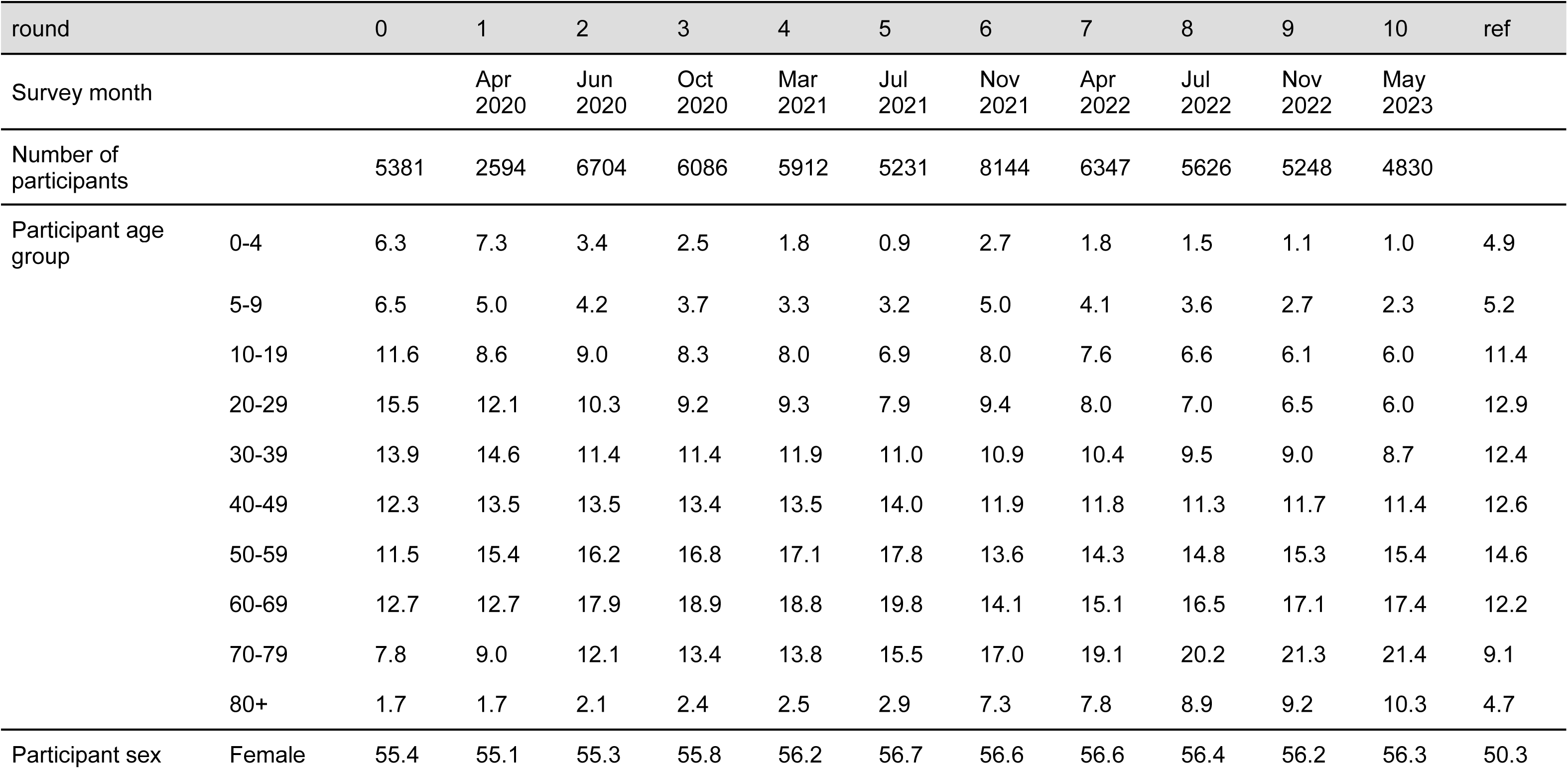
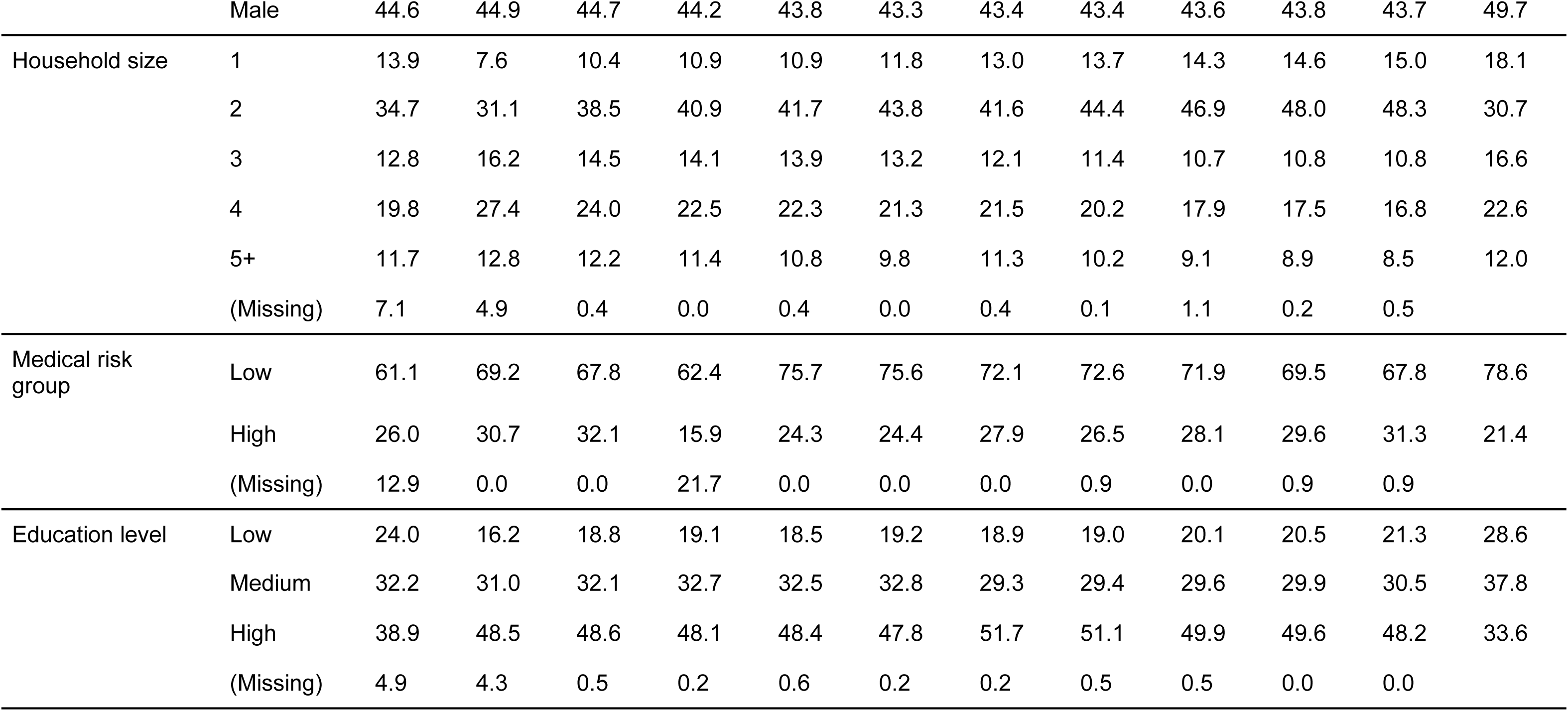
Study population, by survey month, the number of participants per survey round and stratified in percentages by age group, sex, household size, medical risk group and education level. The baseline survey from 2016-2017 is indicated as survey round 0. The final column contains reference percentages for the general population by age group [26], sex [26], household size [31], medical risk status [32], and education level for 15-90 year olds [24]. The Netherlands, February 2016 - November 2022 (n = 62103)

Generally, age groups under 40 years were underrepresented, and age groups between 60 and 80 years were overrepresented compared to the Dutch population [26]. This imbalance seemed to increase over time, suggesting a higher drop-out rate for younger age groups. The study population contained around 56% female participants; a percentage which was constant over all rounds. Participants living in two-person households were overrepresented [31]. This was partly explained by the higher age of the participants; when the household size frequency was corrected for confounding by age and sex, two-person households were less overrepresented, but single-person households remained underrepresented (Suppl S2). The fraction of participants with a high medical risk was higher than in the general population [32], but this was completely explained by confounding with age and sex (Fig. **1**A and Suppl S2). The education level of the participants was higher than in the general population [24] (Fig. **1**B). This overrepresentation became even more pronounced after correcting for age and sex confounding (Suppl S2), because the abundant older participants had on average a lower education level.

**Figure 1:**
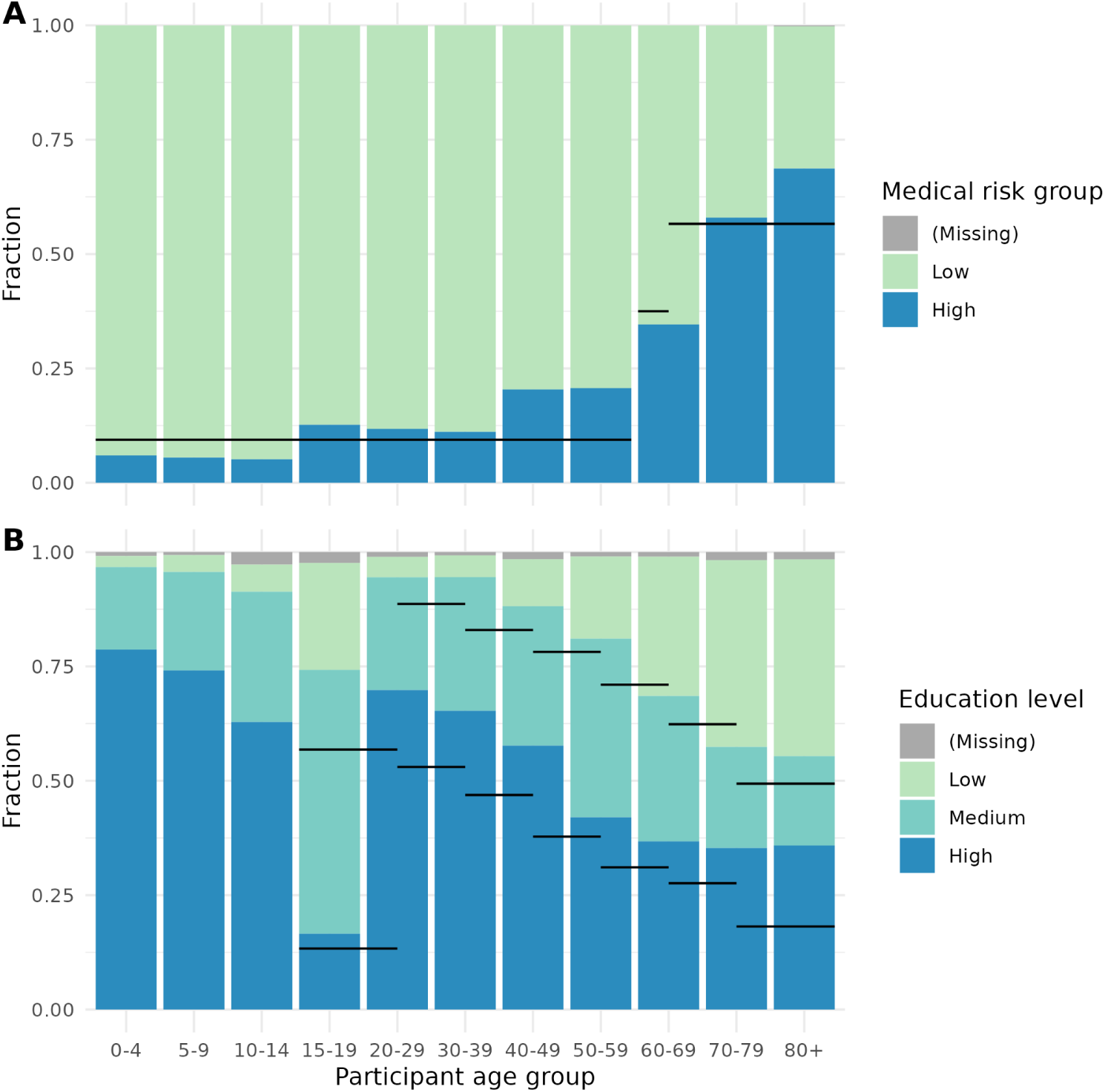
Participant characteristics of the study population by age group. (A) Fraction of participants by medical risk status, for unique participants from PiCo round 4 (March 2021) onwards* (n = 3344). Reference values (black lines) are taken from [32] (B) Fraction of participants by education level, for unique participants. For participants under 15, the highest education level of the parents or guardians is used. For comparability the age group 10-19 is split into 10-14 and 15-19. Reference values (black lines) are taken from [24]. The Netherlands, April 2020 - November 2022 (n = 8211) * From round 4 onwards participants were asked to report current medical conditions, before this round also past medical conditions were reported.

### Number of community contacts

For 1262 questionnaires, the number of community contacts was truncated as they exceeded the maximum of 50 contacts per contact age group. Because of this truncation the 95^th^ percentile of the number of community contacts per participant decreased from 59 to 54.

The number of community contacts in the general population varied over the course of the COVID-19 pandemic (Fig. **2**). Numbers were low in periods with stringent measures and high numbers of hospital admissions. Around the start of 2022 when the Omicron variant became dominant, a short period of lockdown measures did not seem to affect the average number of contacts as much, because 99% of questionnaires in survey round 6 was filled out before these measures came into effect on 19 December 2021. Since the physical distancing measures had been lifted on 25 February 2022, the number of community contacts increased from 13.5 (12.7-14.2) community contacts per person per day in survey round 7 to 15.4 (14.3-16.4) in survey round 10. However, these latest numbers were still 13% lower than the baseline value of 17.8 (17.0-18.5) contacts per day.

**Figure 2:**
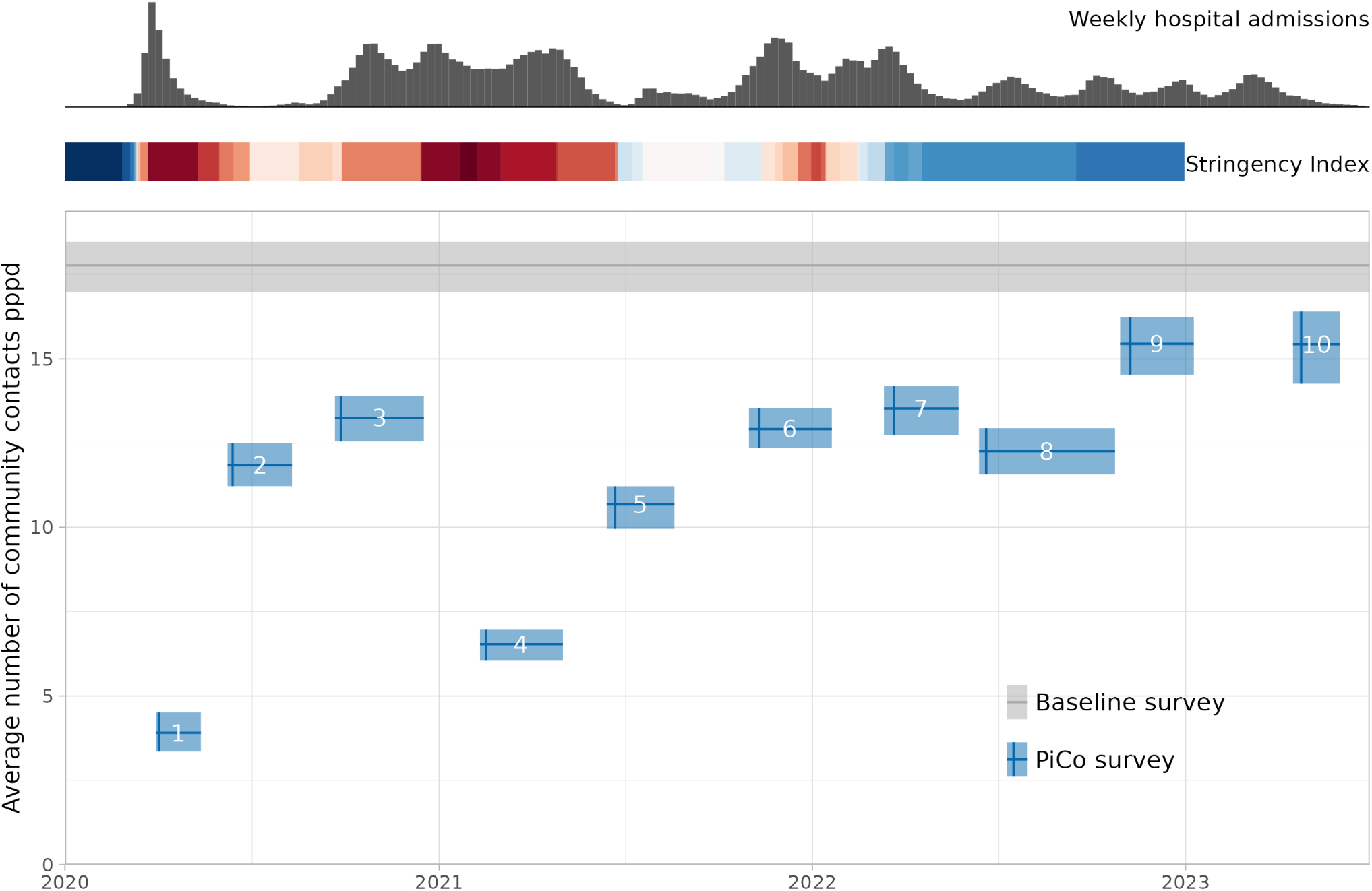
Weighted average number of community contacts per person per day (pppd) in the general population, for the baseline and PiCo survey. Shown is the weighted average number of contacts (horizontal line) and 95% bias-corrected bootstrap interval (shaded area). PiCo rounds are shown from the start to end date, with the median survey date (vertical line) with the survey round number in white. As a timeline reference the weekly number of hospital admissions [40] and the Oxford Stringency Index [41] are depicted on top. The stringency index is color coded from blue (low) to red (high). The Netherlands, April 2020 - November 2022 (n = 62103)

When examining the results in more detail, we observed large differences between age groups (Fig. **3**). In the baseline survey, the number of contacts was highest for the 5-9 age group and gradually decreased with age. During the pandemic, the age groups under 10 years of age were least affected in their contact behaviour. With the exception of the lockdown periods (rounds 1 and 4) that included primary school and day care closures, their number of contacts was around the baseline value. Compared to them, the contacts of 10-19 year olds were more heavily affected, but they have reverted to pre-pandemic levels from 2022 onwards. For all other age groups, the number of contacts were well below baseline during the first two pandemic years. In the last survey rounds, the age groups of 60 years and older approached pre-pandemic behaviour, but the 20-59 year olds have settled at a below-baseline level.

**Figure 3:**
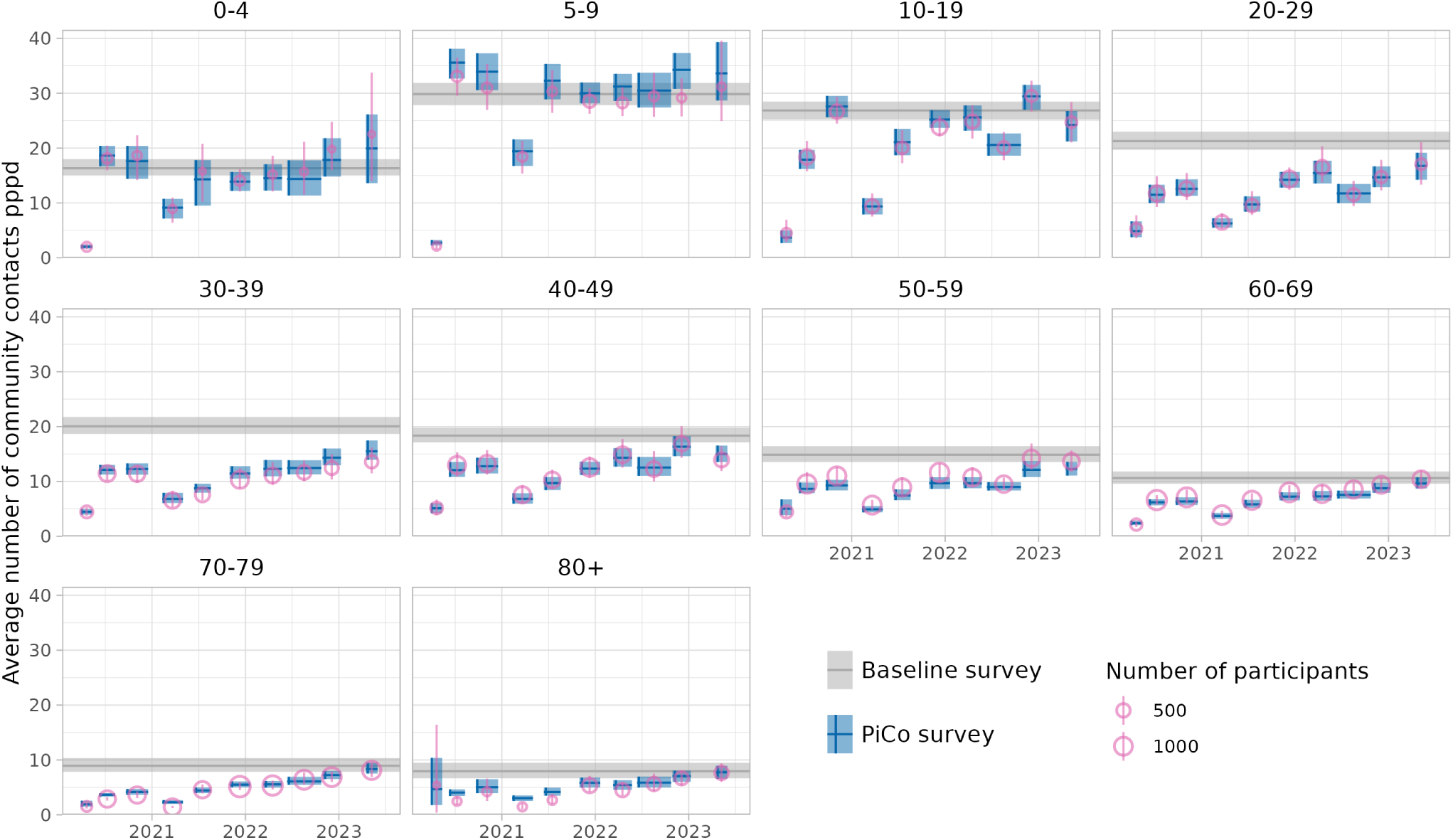
Modelled average number of community contacts per participant per day (pppd) in the general population by age group, for the baseline and PiCo survey. Shown is the average number of contacts (horizontal line) and 95% bias-corrected bootstrap interval (shaded area). PiCo rounds are shown from the start to end date, with the median survey date (vertical line). For the PiCo rounds the reported number of contacts is shown (open circle) with the 95% bias-corrected bootstrap interval (whisker) where the size of the circle is scaled by the number of participants. Reciprocity of contacts is assumed for the modelled number of contacts, not for the reported number of contacts. The Netherlands, April 2020 - November 2022 (n = 62103)

The difference between the modelled (rectangles in Fig. **3**) and reported (circles in Fig. **3**) number of contacts shows how much the reported contacts needed to be adjusted to comply with the assumption of reciprocal contacts. With perfect reporting behaviour, the rectangles and circles would coincide. However, for the 5-9 year olds, for instance, the modelled number of contacts is higher than the reported number of contacts in all survey rounds. This indicates a disbalance where 5-9 year olds reported fewer contacts with other age groups than other age groups with them. Generally, differences between modelled and reported numbers of contacts were smaller than the uncertainty, indicating consistent reporting behaviour.

In the baseline survey the average number of community contacts did not differ by medical risk group (Fig. **4**). Also in the PiCo rounds no difference between the two medical risk groups was observed.

**Figure 4:**
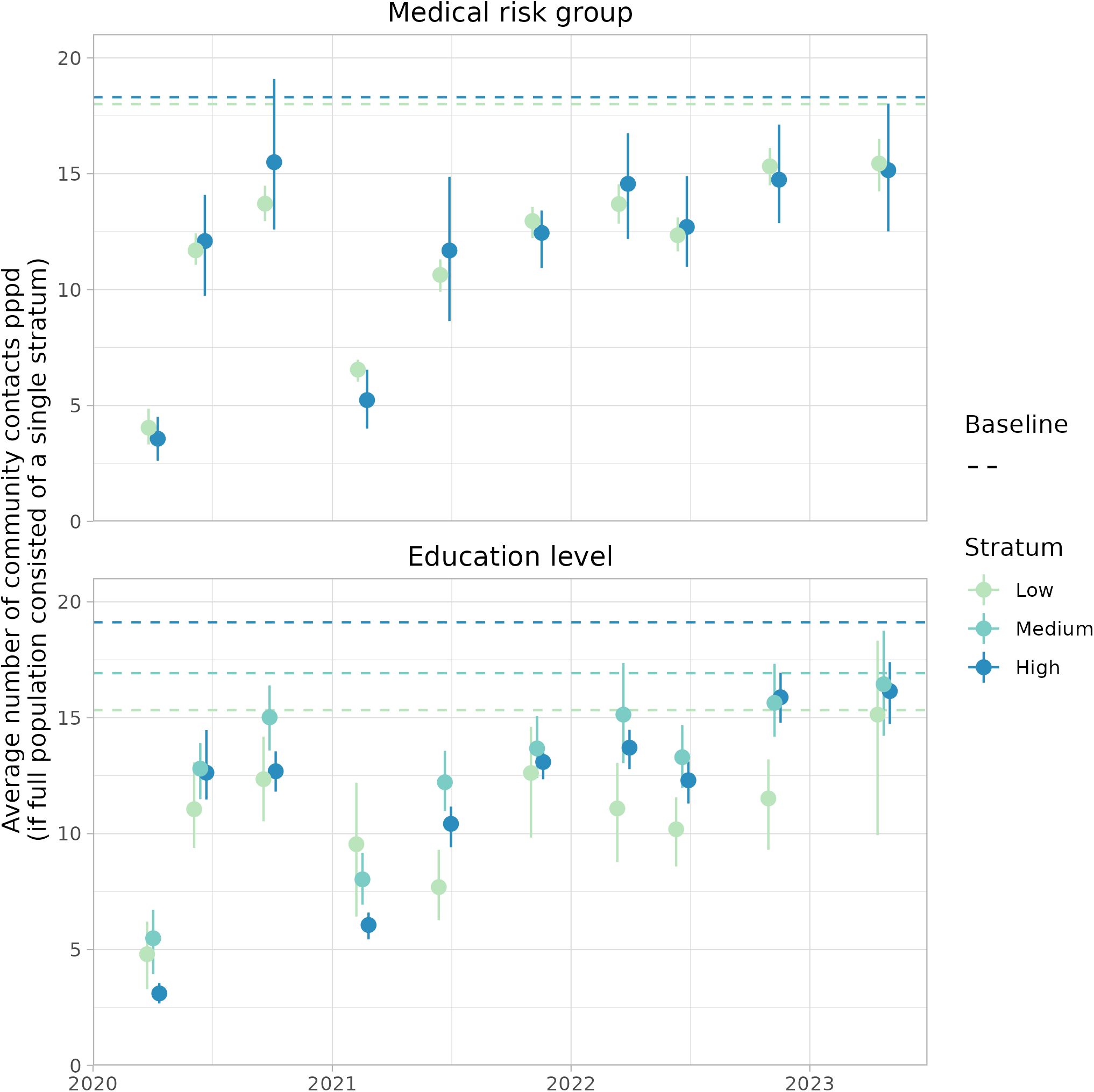
Weighted average number of community contacts per person per day (pppd) by medical risk group (top, n = 59937) and education level (bottom, n = 61565) for the PiCo survey. The averages are calculated by weighting the age groups in each stratum by the age distribution of the general population. Shown is the average number of contacts (point) and 95% bias-corrected bootstrap interval (whisker). For the baseline survey only the average number of contacts is indicated (dashed line). Note that in the upper panel the population is divided in two medical risk strata (high and low). The Netherlands, April 2020 - November 2022

In the baseline survey, the average number of community contacts increased with education level (Fig. **4**). In the PiCo rounds with the most stringent measures (rounds 1 and 4), this ranking was reversed. In all other rounds, with less stringent measures, the average number of community contacts for the low education level is lowest, while middle and high education levels have a similar higher number of contacts.

### Transmission potential

To study the impact of the post-pandemic contact behaviour on the transmission potential of a new respiratory pathogen, we summed the contact matrices of the community and household contacts and assumed values for the relative susceptibility and infectiousness per age group. When all age groups are equally susceptible and infectious, the spectral radius of the resulting matrix is around the baseline value during most of the study period, except for survey rounds 1 and 4 when measures were most stringent (Fig. **5**). When all age groups are equally susceptible, transmission is driven by the age groups with most contacts, which are the youngest age groups that exhibited baseline behaviour during most of the survey rounds. For the early phase of COVID-19, Zhang et al. [2] estimated that younger age groups are less susceptible and older age groups are more susceptible to SARS-CoV-2 infection than adult age groups. When younger age groups are less susceptible, the transmission potential is generally lower than the baseline value, as transmission is now more driven by adult age groups that did structurally decrease their number of contacts. Other reported patterns of the relative susceptibility and infectiousness of COVID-19 lead to similar results (Suppl. S3).

**Figure 5:**
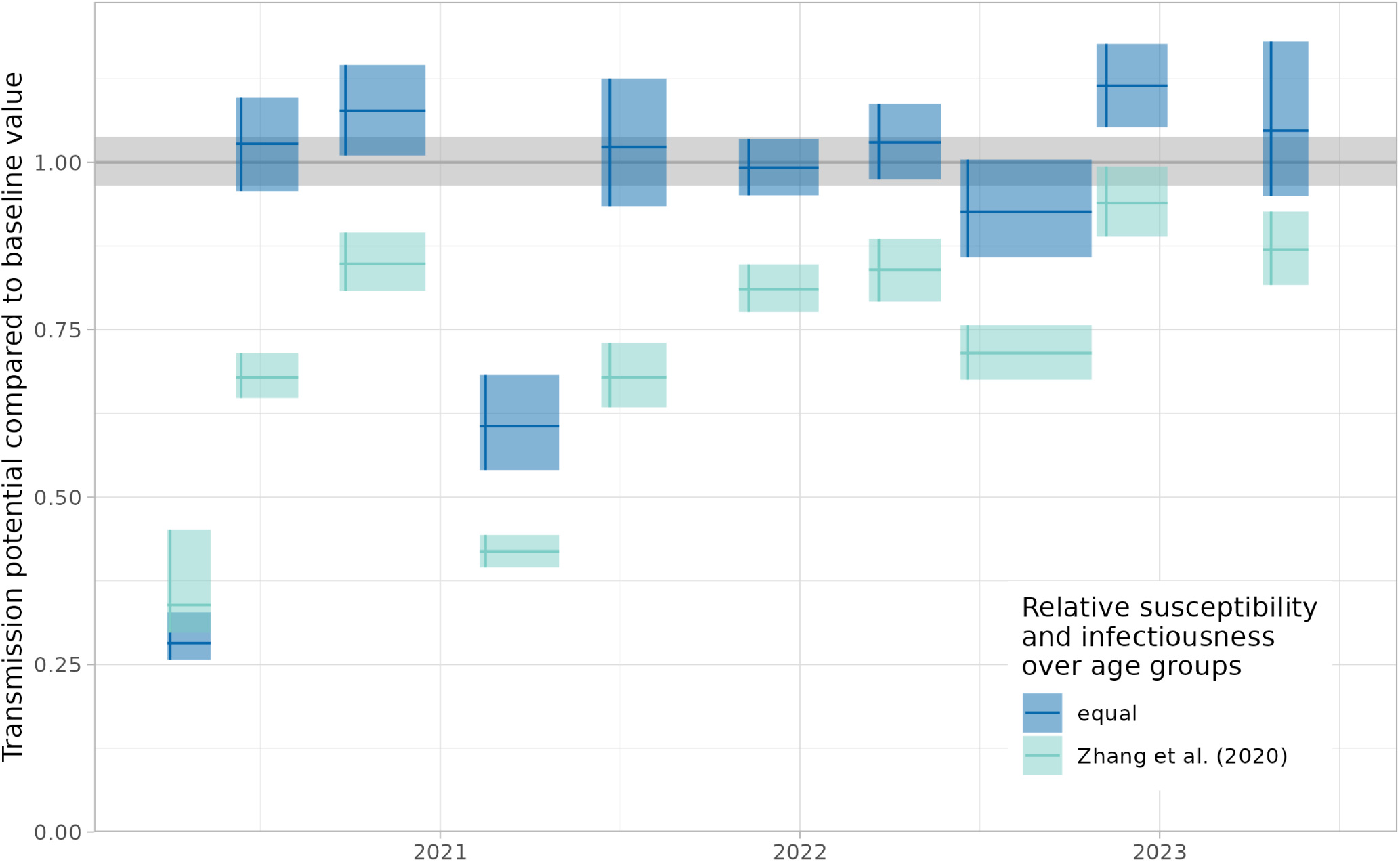
Transmission potential, expressed as the spectral radius of the next generation matrix (NGM) compared to the baseline value, assuming equal relative susceptibility and infectiousness over age groups, or a susceptibility profile for the early phase of COVID-19 [2]. Shown is the ratio (horizontal line) and 95% bias-corrected bootstrap interval (shaded area). PiCo rounds are shown from the start to end date, with the median survey date (vertical line). The Netherlands, April 2020 - November 2022 (n = 62103)

## Discussion

In this study we evaluated whether the Dutch general population has reverted to its pre-pandemic contact behaviour after lifting the COVID-19 measures. Our results show that the population average of community contacts had decreased by 13% in the survey rounds in November 2022 and May 2023 when all control measures had been lifted, compared to the baseline value in 2016-2017. However, children did revert to the high number of contacts they made before the pandemic, retaining their role as potential epidemic drivers. As a consequence, the transmission potential of a newly emerging infection that would spread by a respiratory route mainly depends on how susceptible and infectious children are for this new pathogen.

We found that for children under 20 years and adults over 60 years the number of contacts have reverted to their baseline levels. Only adults in the 20-59 age groups seem to have structurally fewer contacts. A possible explanation is that employees work more hours from home [33] and make fewer contacts while working from home. This trend is further substantiated by the results of the survey question whether participants worked from home last week. For all working participants in age groups 20-69 the fraction (partly) working from home has approximately doubled in the last two survey rounds, compared to a baseline value (Suppl. S4). This is in line with a different survey that reported 37% of employees worked partly from home in 2019, increasing to 45% in 2022 [34].

We found that in the baseline survey the average number of contacts per participant increased with education level. This difference was smaller or even reversed in the periods with lockdown measures (survey rounds 1 and 4), which was also observed in a contact study in Hungary [35]. An explanation is that participants with a lower education level more often have work in essential sectors that preclude working from home [16]. This is confirmed by the results on the survey question whether participants worked from home last week stratified by education level (Suppl. S4). During the lockdown periods around 70% of the highly educated participants worked (partly) from home, against 30% of the participants with a low education level. In the last two survey rounds these percentages dropped to 50% and 25% respectively, which is still higher than the baseline values of 30% and 10% respectively.

We found that the number of contacts did not differ between medical risk groups (i.e. indication for influenza vaccination) in any of the survey rounds nor the baseline survey. This was confirmed by the COVIMOD study in Germany [14] that used the same medical risk definition. However, in some study rounds of the CONNECT study in Canada, it was found that participants with comorbidities had fewer contacts than participants without comorbidities [13]. This would suggest that the absence of an association between number of contacts and medical risk group could be caused by our broad risk definition; persons with a very high medical risk may well have reduced their number of contacts. Participants with a high medical risk could also have had safer contacts with reduced risk of transmission by keeping distance and/or wearing protection. However, for these types of contacts we found no difference between medical risk groups (Suppl. S5).

Our finding that the different contact patterns have a limited impact on the spread of a new pathogen if all age groups are equally susceptible is in apparent contrast with the findings reported in the final CoMix round held in November 2022 in four European countries [36]. For the Netherlands, an average number of 9.9 (9.0-10.8) contacts per person per day was found, which is lower than the 15.4 (14.5-16.2) contacts per person per day that we found in November 2022 (PiCo round 9). Using an earlier survey from 2006-2007 as baseline [37], the CoMix study reported that the transmission potential in the Netherlands had decreased by 20%, while our study suggests no change in transmission potential when assuming equal susceptibility and infectiousness over age groups. Possible explanations could be found in the different design, study population and study period of the baseline survey, and in the fatigue effect observed for the Dutch CoMix study which leads participants to report decreasing numbers of contacts due to the high frequency of reporting [6,38].

The strengths of this study include the identical study design of the baseline and PiCo surveys, which enables the comparison of current to past contact behaviour. Both surveys are conducted with participants of all age groups, which is essential to study the spread in the entire population. The CONNECT study in Canada has a similar study design [9]; a post-pandemic survey of this study would be valuable to compare our main results to. Another strength is that the contact questionnaire is embedded in a larger serosurveillance study, providing a wealth of additional information on the participants. This allowed us to stratify our analysis by medical risk group and education level.

Limitations include the possibility that the study population was on average more compliant to COVID-19 measures than the general population. Evidence for this is for instance their higher COVID-19 vaccine uptake [22]. Also, participants may have reported contacts differently compared to the baseline survey, because they have been made more aware of their contacts by COVID-19 information campaigns. Finally, we have weighted participants by age and sex, but not by education level due to the lack of reference values for participants younger than 15 years. Since participants with a high education level are overrepresented, and tend to have a higher number of contacts than average with relaxed measures and a lower number of contacts than average with stringent measures, a consequence could be that the numbers of contacts are overestimated in periods with relaxed measures and underestimated in periods with stringent measures.

Contact surveys have proven invaluable during the COVID-19 pandemic in monitoring changes in contact behaviour due to evolving measures or compliance with them. The baseline and PiCo surveys have provided direct quantitative evidence to inform tailored infection control policies in the Netherlands. Continuation of such contact surveys is therefore essential, even in a non-pandemic setting, not only to signal changes in contact patterns that affect the transmission potential, but also to provide a baseline for the next pandemic.

## Declarations

### Ethics approval and consent

This study was approved by the Medical Ethics Committee (MEC-U), the Netherlands (Clinical Trial Registration NTR8473), conformed to the principles embodied in the Declaration of Helsinki, and all participants provided written informed consent.

### Availability of data and materials

The cleaned data is published online at https://doi.org/10.5281/zenodo.10370353 [25].

All code for data cleaning and analysis is publicly available at https://github.com/rivm-syso/pico_contactsurvey [39].

### Authors’ contributions

F.v.d.K, G.d.H, H.d.M. and J.W. designed the study, F.v.d.K, G.d.H, C.v.d.H and E.V. collected the data, J.B. did the analyses, J.B. wrote the main manuscript text. All authors reviewed the manuscript.

## Data Availability

The cleaned data are available online at

https://doi.org/10.5281/zenodo.10370353

## Acknowledgments

We gratefully acknowledge the participants of the PIENTER3 and PIENTER Corona (PICO) studies. We would also like to thank Federica Giardina (Radboud University Medical Center), Ganna Rozhnova (Utrecht University) and Otilia Boldea (Tilburg University) for their valuable input.

