## Supplementary material for "Contact behaviour before, during and after the COVID-19 pandemic in the Netherlands: evidence from contact surveys in 2016-2017 and 2020-2023"

S1 Study design and data

S2 Study population weighted by age group and sex

S3 Sensitivity analysis for transmission potential

S4 Working from home

S5 Contact type by medical risk group

### S1 Study design and data

Details on the study design and selection of participants were described in publications on the baseline survey [1] and the PiCo surveys [2–4]. Here we will briefly summarise aspects that are relevant to our study.

#### Participant selection

For the baseline survey, participants were randomly selected from the national Personal Records Database (BRP) [5] via a two-stage cluster design, comprising 40 municipalities in five regions nationwide [1]. The national sample consisted of 5745 participants (response rate of 14.4%). The participants who consented to be approached for follow-up and could be contacted (i.e. 80% of the baseline survey participants) were invited for the first round of the PiCo survey in April 2020, 53% of whom participated [2]. For the second PiCo round (June 2020), 26,854 additional participants were randomly selected from the BRP from all regions of the Netherlands, 16.7% of whom participated [3]. The participants of PiCo rounds 1 and 2 were reinvited for all subsequent rounds. For the sixth PiCo round (November 2021), 65,690 additional participants were randomly selected from the BRP from all regions of the Netherlands, 4.8% of whom participated [4]. Also these new participants of PiCo rounds 6 were reinvited for all subsequent rounds. The participation rate for each of these groups of participants is shown in Table **1**. Note that these are lower bounds as participants may have requested to be removed from the study or may have passed away over the course of the survey. In Table **2** the median, start and end date for each round are listed. These dates are used in Figures 2, 3, and 5 in the main text.

**Table 1:** Participation by groups (G) that were invited for PiCo rounds 1, 2 and 6, and overall participation. Columns show the number of participants (n) and participation rates (%) .

| Round | G1 (n) | G1 (%) | G2 (n) | G2 (%) | G6 (n) | G6 (%) | Overall (n) | Overall (%) |
|---|---|---|---|---|---|---|---|---|
| 1 | 2,594 | 100.0 | | | | | 2,594 | 100.0 |
| 2 | 2,207 | 85.1 | 4,497 | 97.2 | | | 6,704 | 92.8 |
| 3 | 2,037 | 78.5 | 4,049 | 87.5 | | | 6,086 | 84.3 |
| 4 | 2,036 | 78.5 | 3,876 | 83.8 | | | 5,912 | 81.9 |
| 5 | 1,824 | 70.3 | 3,407 | 73.6 | | | 5,231 | 72.4 |
| 6 | 1,691 | 65.2 | 3,167 | 68.4 | 3,286 | 98.9 | 8,144 | 77.2 |
| 7 | 1,355 | 52.2 | 2,527 | 54.6 | 2,465 | 74.2 | 6,347 | 60.2 |
| 8 | 1,234 | 47.6 | 2,307 | 49.8 | 2,085 | 62.7 | 5,626 | 53.4 |
| 9 | 1,193 | 46.0 | 2,187 | 47.3 | 1,868 | 56.2 | 5,248 | 49.8 |
| 10 | 1,126 | 43.4 | 1,992 | 43.0 | 1,712 | 51.5 | 4,830 | 45.8 |

**Table 2:** Time frame of survey rounds of baseline survey (round 0 ) and PiCo survey rounds. Shown are median, start and end date for each round.

| Round | Median date | Start date | End date |
|---|---|---|---|
| 0 | 2017-01-29 | 2016-01-10 | 2017-12-03 |
| 1 | 2020-04-02 | 2020-03-30 | 2020-05-13 |
| 2 | 2020-06-13 | 2020-06-08 | 2020-08-10 |
| 3 | 2020-09-27 | 2020-09-21 | 2020-12-17 |
| 4 | 2021-02-16 | 2021-02-10 | 2021-05-02 |
| 5 | 2021-06-22 | 2021-06-14 | 2021-08-19 |
| 6 | 2021-11-10 | 2021-10-31 | 2022-01-20 |
| 7 | 2022-03-22 | 2022-03-12 | 2022-05-24 |
| 8 | 2022-06-20 | 2022-06-13 | 2022-10-24 |
| 9 | 2022-11-08 | 2022-10-29 | 2023-01-09 |
| 10 | 2023-04-24 | 2023-04-16 | 2023-06-01 |

### Changes in survey questions

Most questions on participant and contact characteristics remained the same over the course of all rounds, including the baseline survey. Some survey questions however changed during the study period, summarised in Table **3**.

Contacts' ages were reported in age groups: 0–4, 5–9, 10–19, 20–29, 30–39, 40–49, 50–59, 60–69, 70–79, 80–89 and ≥ 90, but from PiCo round 3 onwards the contact age group 10-19 was subdivided in 10-14 and 15-19. Participants could further distinguish their contacts, but this distinction changed from round to round. In the baseline survey and Pico rounds 1 and 8, the number of men and women in each contact age group was reported. In PiCo rounds 2, 3, and 10 this distinction was replaced by whether contacts took place within or outside 1.5 meters. In all other PiCo rounds, the category for contacts within 1.5 meters, was further subdivided in whether the contact occurred with or without personal protective equipment, such as a face mask.

A question about working from home was included in the PiCo survey. To obtain a baseline value, participants of rounds 1 and 2 were also asked about working from home in the period preceding the COVID-19 pandemic.

**Table 3:** Survey questions that have changed during the study period of the baseline survey (indicated as round 0) and the PiCo survey (rounds 1 to 10). The close/distant distinction is defined as less/more than 1.5 m apart. An 'x' denotes which version of the survey question was posed in that round.

| Survey question | | 0 | 1 | 2 | 3 | 4 | 5 | 6 | 7 | 8 | 9 | 10 |
|---|---|---|---|---|---|---|---|---|---|---|---|---|
| Household composition | including participant | x | x | | | | | | | | | |
| | excluding participant | | | x | x | x | x | x | x | x | x | x |
| Medical conditions | currently or in past | x | x | x | | | | | | | | |
| | currently | | | | | x | x | x | x | x | x | x |
| Working from home | before COVID-19 | | x | x | | | | | | | | |
| | last week | | x | x | x | x | x | x | x | x | x | x |
| Check on contacts | | | x | x | x | x | x | x | x | x | x | x |
| Contact age group 10-19 | 10-19 | x | x | x | | | | | | | | |
| | 10-14 and 15-19 | | | | x | x | x | x | x | x | x | x |
| Contact distinction | men / women | x | x | | | | | | | x | | |
| | close / distant | | | x | x | | | | | | | x |
| | close with/out protection / distant | | | | | x | x | x | x | | x | |

### Contact question

The contact question as was asked in PiCo round 7 is shown in Figure **1**. The check on the number of contacts (Did you have contacts with other persons yesterday?) was only present in the Pico surveys, not in the baseline survey. The distinctions to be made between contacts could differ by PiCo round (see Tab. **3**).

10. How many persons did you have contact with yesterday (excluding household members)?

*A contact happens when you, for instance, talk to someone, touch someone, kiss someone or play sports with someone. Also remember contacts at work or school. Telephone conversations, e-mail and other online contacts should not be counted.*

*Please distinguish between <u>protected</u> and <u>unprotected</u> contacts that happened within 1.5 meters and contacts that happened at a distance of 1.5 m or more. Protected contacts are, for instance, when both persons (you and the person you have contact with) wear a face mask or face shield or when a plastic barrier is placed between both persons (such as at the cashier in shops).*

*Indicate this by age group.*

*This is a difficult question; we'd like to emphasize that an approximate answer suffices.*

Did you have contacts with other persons yesterday (<u>excluding household members</u>)?

☐ Yes, fill out the table below (you can leave cells empty)

☐ No ➔ **continue with question 11**

*NB Indicate in NUMBERS how many contacts you had in the tabel below. Do not use crosses or other symbols to indicate the number of contacts.*

| **Age group** | **Number of <u>unprotected</u> contacts WITHIN 1,5 meters** | **Number of <u>protected</u> contacts WITHIN 1,5 meters** | **Number of contacts at a distance of 1.5 meters or more** |
|---|---|---|---|
| 0 - 4 year | | | |
| 5 - 9 year | | | |
| 10 - 14 year | | | |
| 15 - 19 year | | | |
| 20 - 29 year | | | |
| 30 - 39 year | | | |
| 40 - 49 year | | | |
| 50 - 59 year | | | |
| 60 - 69 year | | | |
| 70 - 79 year | | | |
| 80 - 89 year | | | |
| 90+ year | | | |

11. Yesterday was a:

☐ Monday ☐ Tuesday ☐ Wednesday ☐ Thursday ☐ Friday ☐ Saturday ☐ Sunday

**Figure 1:** Contact question of PiCo round 7 (translated from Dutch).

### Valid household composition

Participants were asked about their household size and composition, i.e. the age and sex of all household members. In the baseline survey and round 1 of the PiCo survey, the participants should include themselves, but from round 2 of the PiCo survey they should exclude themselves. As a consequence, from round 2 onwards the household composition could be empty, either because the participant lived in a single-person household or because they skipped the question. All non-empty household compositions were indicated to be valid, as well as empty household compositions of participants who reported to live in a single-person household from round 2 onwards. To harmonize the different surveys, persons with the same age and sex as the participant were deleted from the reported household composition in the baseline survey and round 1 of the PiCo survey. Participants without a valid household composition are only excluded for the analysis of contacts with household members, but they are included for the analysis of community contacts.

### Medical risk status

In the baseline survey and PiCo rounds 1 and 2, the participant was asked about current and previous medical conditions. From PiCo round 4 onwards, the question was restricted to current medical conditions. In PiCo round 3 no medical questions were posed.

Using these medical conditions, the medical risk status of each participant was based on whether they would be indicated for influenza vaccination. Following the current guidelines [6] these conditions include diabetes (any kind), respiratory disease, liver disease, immunocompromised condition, cancer, asplenia, renal disease, cardiovascular disease, neurological condition, transplant patients, and/or morbid obesity (Body Mass Index ≥ 40, calculated from the length and weight of the participant). When at least one of these conditions applied, the participant was classified in the high medical risk category. When none of the medical condition questions were answered (or posed), the medical risk status was imputed. Because the medical risk questions changed around PiCo round 3, we imputed the missing medical risk status for the periods before round 3 and from round 3 onwards separately. For each period, it was determined whether a participant had an unambiguous risk status, i.e., only high or only low, apart from the missing data. If so, the missing risk status was replaced by the unambiguous risk status. For instance, a participant with a medical risk status of high, missing and missing in round 0, 1 and 2 would have an unambiguous high risk status, and the missing risk status would be replaced by high medical risk. On the other hand, a participant with a medical risk status of high, low and missing in round 0, 1 and 2 would have an ambiguous risk status, and the missing risk status is not imputed.

### Valid contact data

Only participants who provided valid contact data are included in the data. In the PiCo survey, participants were first asked whether they had had any contacts outside the household on the previous day, before filling out the actual number of contacts. Participants who answered ‘yes’ and reported a number of contacts, and participants who answered ‘no’ and reported zero contacts were included. Participants who answered ‘yes’ but did not report any contacts, and participants who answered ‘no’ but did report a number of contacts were excluded. Participants who did not answer the check question were only included if they provided a number of contacts. Participants who did not answer the check question and did not provide any contacts were excluded, presuming they skipped the question.

The baseline survey lacked the question to check whether a participant had any contacts on the previous day. Instead, we used the question on which day it was yesterday (question 11 in **1**) as a check. Participants who filled out this questions were included, with and without reported contacts. Participants who did not provide the contact day were only included if they reported a number of contacts. Participants who did not provide the contact day and did not report any contacts were excluded, presuming they skipped the question.

### S2 Study population corrected for confounding by age group and sex

**Table 4:** Study population characteristics corrected for confounding, using the age group and sex of the general population [5]. The survey month, the number of participants per survey round and stratified in percentages by household size, medical risk group and education level. The baseline survey from 2016-2017 is indicated as survey round 0. Before weighting the missing values were omitted. The final column contains reference percentages for the household size [7], medical risk group [8], and education level for 15-90 year olds [6].

| round | | 0 | 1 | 2 | 3 | 4 | 5 | 6 | 7 | 8 | 9 | 10 | ref |
|---|---|---|---|---|---|---|---|---|---|---|---|---|---|
| Survey month | | | Apr 2020 | Jun 2020 | Oct 2020 | Mar 2021 | Jul 2021 | Nov 2021 | Apr 2022 | Jul 2022 | Nov 2022 | May 2023 | |
| Number of participants | | 5381 | 2594 | 6704 | 6086 | 5912 | 5231 | 8144 | 6347 | 5626 | 5248 | 4830 | |
| Household size | 1 | 22.0 | 13.5 | 11.3 | 11.0 | 11.2 | 11.4 | 11.9 | 12.1 | 13.0 | 11.9 | 12.2 | 18.1 |
| | 2 | 34.9 | 31.6 | 33.3 | 33.9 | 34.2 | 34.4 | 34.6 | 35.1 | 35.6 | 35.1 | 35.2 | 30.7 |
| | 3 | 12.4 | 15.6 | 14.6 | 14.6 | 14.1 | 13.6 | 13.8 | 13.4 | 13.1 | 13.3 | 13.4 | 16.6 |
| | 4 | 19.5 | 26.9 | 26.6 | 26.1 | 26.3 | 26.9 | 25.8 | 25.9 | 24.9 | 25.4 | 24.8 | 22.6 |
| | 5+ | 11.2 | 12.5 | 14.2 | 14.4 | 14.2 | 13.7 | 13.9 | 13.5 | 13.4 | 14.2 | 14.3 | 12.0 |
| Medical risk group | Low | 71.1 | 69.3 | 70.7 | 81.7 | 79.5 | 80.7 | 77.4 | 79.5 | 79.9 | 79.3 | 78.4 | 78.6 |
| | High | 28.9 | 30.7 | 29.3 | 18.3 | 20.5 | 19.3 | 22.6 | 20.5 | 20.1 | 20.7 | 21.6 | 21.4 |
| Education level | Low | 30.8 | 22.6 | 20.3 | 19.4 | 18.6 | 18.2 | 18.0 | 16.9 | 17.0 | 16.7 | 16.8 | 28.6 |
| | Medium | 33.5 | 34.4 | 32.3 | 32.9 | 32.9 | 33.1 | 31.8 | 31.6 | 31.5 | 31.8 | 32.2 | 37.8 |
| | High | 35.7 | 43.0 | 47.4 | 47.7 | 48.5 | 48.7 | 50.3 | 51.5 | 51.6 | 51.5 | 51.1 | 33.6 |

### S3 Sensitivity analysis for transmission potential

To assess how sensitive the transmission potential is to assumptions on the relative susceptibility and infectiousness by age group for COVID-19, we tried several parameterisations from literature. Zhang et al. [7] estimated susceptibility by age group, while keeping infectiousness constant. Franco et al. [8] used an NGM approach fixing either infectiousness and estimating susceptibility (scenario A) or vice versa (scenario B). Finally, Klinkenberg et al. [9] assumed susceptibility and infectiousness were varying by age group but identical. Although these assumptions lead to slightly different estimates for the transmission potential, they all are distinctly different from the assumption that susceptibility and infectiousness are equal for all age groups (Fig. **2**).

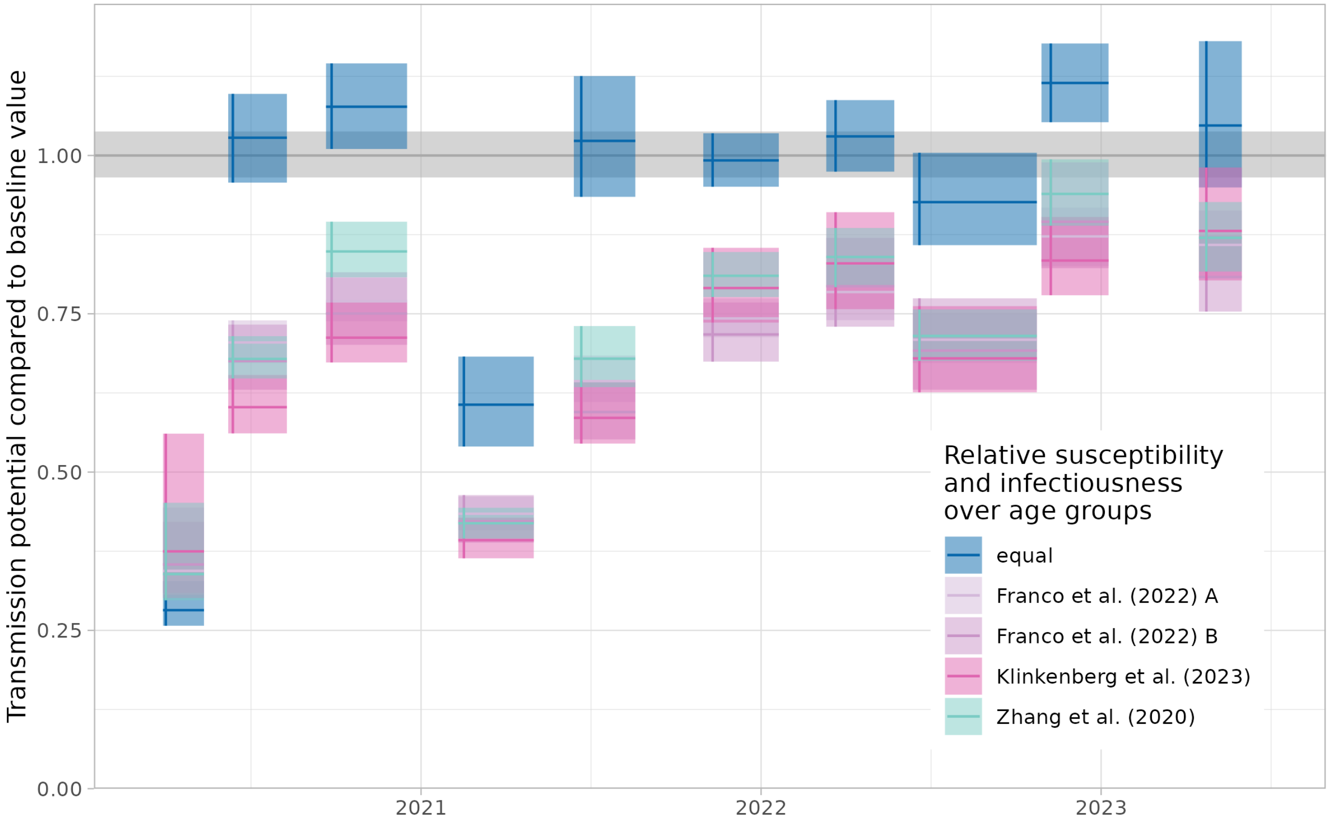


**Figure 2:** Transmission potential, expressed as the spectral radius of the next generation matrix (NGM) compared to the baseline value, for different assumptions of relative susceptibility and infectiousness over age groups. Shown is the ratio (horizontal line) and 95% bias-corrected bootstrap interval (shaded area). PiCo rounds are shown from the start to end date, with the median survey date (vertical line).

**Table 5:** Assumptions for relative susceptibility and infectiousness by age group, for sensitivity analysis of transmission potential.

| reference | type | 0-4 | 5-9 | 10-19 | 20-29 | 30-39 | 40-49 | 50-59 | 60-69 | 70-79 | 80+ |
|---|---|---|---|---|---|---|---|---|---|---|---|
| equal | sus | 1.000 | 1.000 | 1.000 | 1.00 | 1.000 | 1.000 | 1.00 | 1.000 | 1.000 | 1.000 |
| | inf | 1.000 | 1.000 | 1.000 | 1.00 | 1.000 | 1.000 | 1.00 | 1.000 | 1.000 | 1.000 |
| Franco et al. (2022) A | sus | 0.182 | 0.550 | 0.603 | 1.00 | 1.172 | 1.009 | 0.88 | 0.869 | 0.846 | 0.805 |
| | inf | 0.540 | 0.550 | 0.560 | 0.59 | 0.700 | 0.760 | 0.90 | 0.990 | 0.990 | 0.990 |
| Franco et al. (2022) B | sus | 0.400 | 0.390 | 0.380 | 0.79 | 0.860 | 0.800 | 0.82 | 0.880 | 0.740 | 0.740 |
| | inf | 0.346 | 0.892 | 1.310 | 1.00 | 0.645 | 3.783 | 1.32 | 0.266 | 1.277 | 0.099 |
| Klinkenberg et al. (2023) | sus | 1.000 | 1.000 | 3.050 | 5.75 | 3.540 | 3.710 | 4.36 | 5.690 | 5.320 | 7.210 |
| | inf | 1.000 | 1.000 | 3.050 | 5.75 | 3.540 | 3.710 | 4.36 | 5.690 | 5.320 | 7.210 |
| Zhang et al. (2020) | sus | 0.340 | 0.340 | 0.670 | 1.00 | 1.000 | 1.000 | 1.00 | 1.235 | 1.470 | 1.470 |
| | inf | 1.000 | 1.000 | 1.000 | 1.00 | 1.000 | 1.000 | 1.00 | 1.000 | 1.000 | 1.000 |

### S4 Working from home

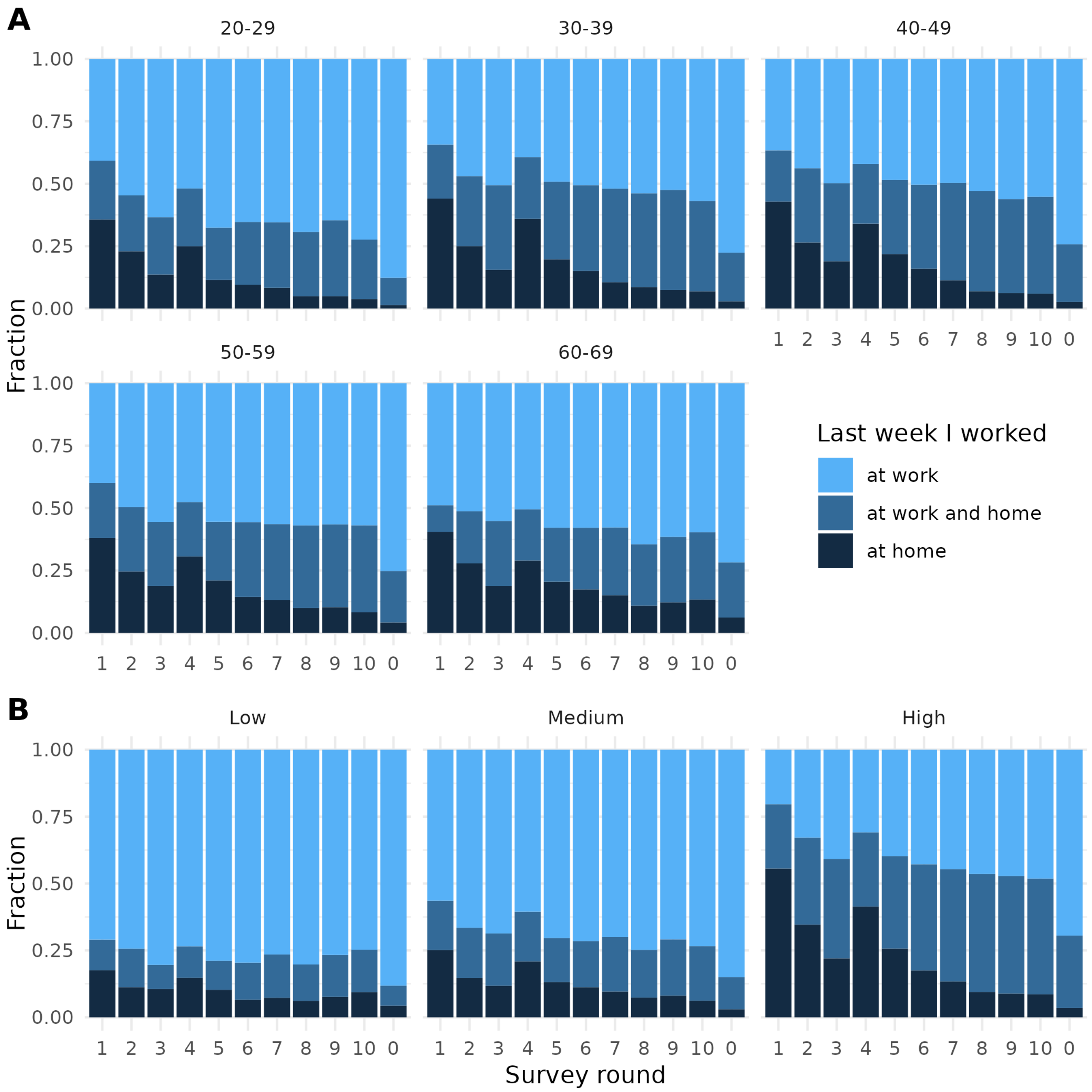


**Figure 3:** Fraction of participants with employment in working ages 20-69 that (partly) worked from home in the previous week, by (A) age group and (B) education level. The baseline value (at round 0, for ease of comparison plotted next to the most recent rounds) is the fraction of participants who (partly) worked from home in the pre-COVID-19 period, answered by participants in rounds 1 and 2.

### S5 Contact type by medical risk group

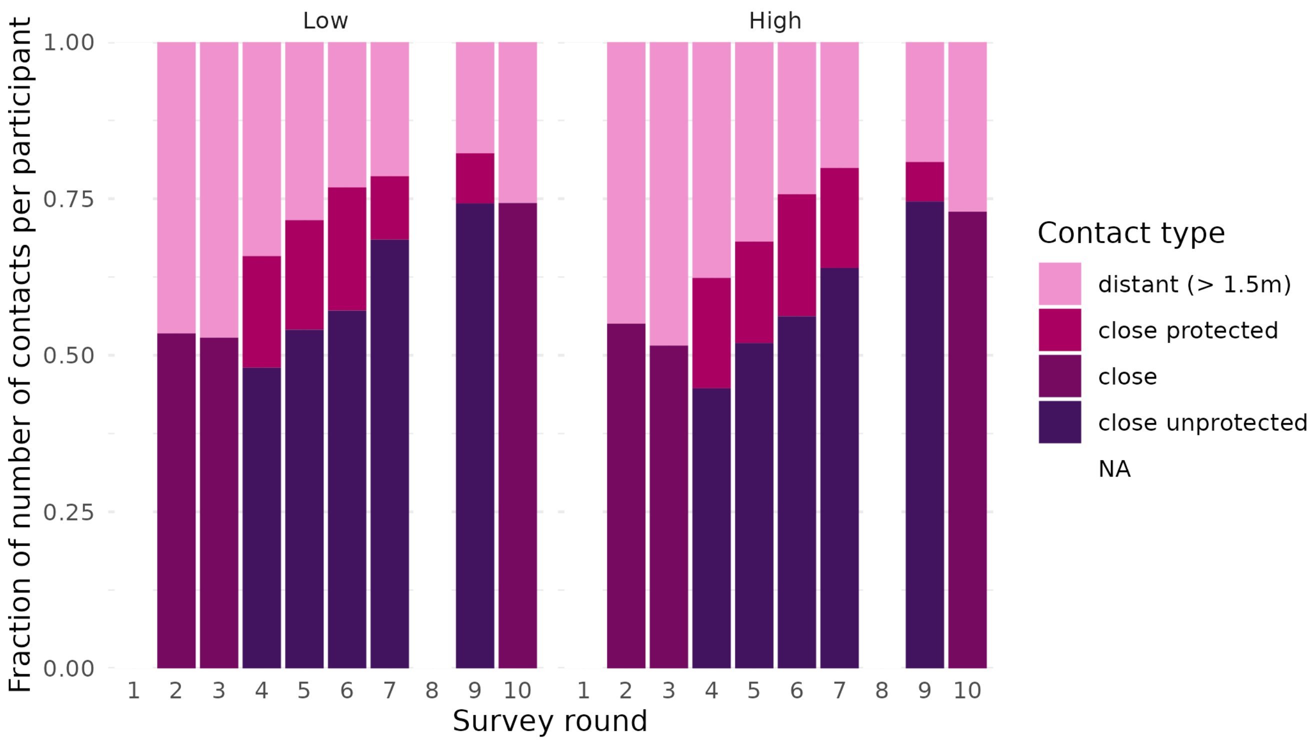


**Figure 4:** Fraction of contacts per participant, stratified by contact type: distant (more than 1.5 m), close (less than 1.5 m), close unprotected (less than 1.5 m without protection), close protected (less than 1.5 m with protection) by survey round. In rounds 1 and 8 contacts were stratified by gender. Fractions are weighted by age group distribution of the general population.
